# Novel deep learning algorithm predicts the status of molecular pathways and key mutations in colorectal cancer from routine histology images

**DOI:** 10.1101/2021.01.19.21250122

**Authors:** Mohsin Bilal, Shan E Ahmed Raza, Ayesha Azam, Simon Graham, Muhammad Ilyas, Ian A. Cree, David Snead, Fayyaz Minhas, Nasir M. Rajpoot

**Affiliations:** Department of Computer Science, University of Warwick, UK; University Hospitals Coventry and Warwickshire, UK; Faculty of Medicine & Health Sciences, University of Nottingham, UK; International Agency for Research on Cancer (AIRC), Lyon, France

## Abstract

**Background:** Determining molecular pathways involved in the development of colorectal cancer (CRC) and knowing the status of key mutations are crucial for deciding optimal target therapy. The goal of this study is to explore machine learning to predict the status of the three main CRC molecular pathways – microsatellite instability (MSI), chromosomal instability (CIN), CpG island methylator phenotype (CIMP) – and to detect BRAF and TP53 mutations as well as to predict hypermutated (HM) CRC tumors from whole-slide images (WSIs) of colorectal cancer (CRC) slides stained with Hematoxylin and Eosin (H&E).

**Methods:** We propose a novel iterative draw-and-rank sampling (IDaRS) algorithm to select representative sub-images or tiles from a WSI given a single WSI-level label, without needing any detailed annotations at the cell or region levels. IDaRS is used to train a deep convolutional network for predicting key molecular parameters in CRC (in particular, prediction of HM tumors and the status of three main CRC molecular pathways – MSI, CIN, CIMP – as well as the detection of two key mutations, BRAF and TP53) from digitized images of routine H&E stained tissue slides of CRC patients (n=497 for TCGA cohort and n=47 cases for the Pathology AI Platform or PAIP cohort). Visual fields most predictive of each pathway and HM tumors identified by IDaRS are analyzed for verification of known histological features for the first time to reveal novel histological features. This is achieved by systematic, data-driven analysis of the cellular composition of strongly predictive tiles.

**Findings:** IDaRS yields high prediction accuracy for prediction of the three main CRC genetic pathways and key mutations by deep learning based analysis of the WSIs of H&E stained slides. It achieves the state-of-the-art AUROC values of 0.90, 0.83, and 0.81 for prediction of the status of MSI, CIN, and HM tumors for the TCGA cohort, which is significantly higher than any other currently published methods on that cohort. We also report prediction of status of CIMP pathway (CIMP-High and CIMP-Low) from H&E slides, with an AUROC of 0.79. We analyzed key discriminative histological features associated with HM tumors and each molecular pathway in a data-driven manner, via an automated quantitative analysis of the cellular composition of tiles strongly predictive of the corresponding molecular status. A key feature of the proposed method is that it enables a systematic and data-driven analysis of the cellular composition of image tiles strongly predictive of the various molecular parameters. We found that relatively high proportion of tumor infiltrating lymphocytes and necrosis are found to be strongly associated with HM and MSI, and moderately associated with CIMP-H and genome-stable (GS) cases, whereas relatively high proportions of neoplastic epithelial type 2 (NEP2), mesenchymal and neoplastic epithelial type 1 (NEP1) cells are found to be associated with CIN cases.

**Interpretation:** Automated prediction of genetic pathways and key mutations from image analysis of simple H&E stained sections with a high accuracy can provide time and cost-effective decision support. This work shows that a deep learning algorithm can mine both visually recognizable as well as sub-visual histological patterns associated with molecular pathways and key mutations in CRC in a data-driven manner.

**Funding:** This study was funded by the UK Medical Research Council (award MR/P015476/1).

## Introduction

Molecular pathways of colorectal cancer (CRC) carcinogenesis can help to explain the basis for diversity in disease progression and growth in different patients.^1^ For instance, the chromosomal instability (CIN) pathway is associated with lower rates of overall and progression-free survival in CRC^2^ whereas microsatellite instability (MSI) can stratify a group of patients as likely respondents to immunotherapy.^3–6^ Further classification is possible considering different subgroups of CpG island methylator phenotype (CIMP) and MSI^7^ based on different responses to adjuvant therapy and survival. A recent survey lists several studies showing different classifications based on these pathways and their clinical impacts.^3^ There is increasing evidence evaluating shared and distinguished molecular characteristics and pathways for CRC tumor subtyping^1,8^ and establishing the association of these pathways with patient prognosis, overall survival and response to specific treatments, particularly targeted therapy.^3,9,10^ For instance, Pembrolizumab has recently been approved by the US Food and Drug Authority (FDA) for the first-line treatment of patients with MSI or mismatch repair deficient (dMMR) colorectal cancer by immunotherapy.^11^

The standard genetic (e.g., polymerase chain reaction or PCR) and immunohistochemistry (IHC) testing may incur time delays and additional costs.^3,4^ In addition, molecular testing assays often require tissue consumption which may be quite restrictive due to limited tissue availability (such as diagnostic biopsies).^12^ On the other hand, visual examination of Hematoxylin and Eosin (H&E) stained tissue slides remains the ‘gold standard’ for diagnosis of CRC. And with recent surge in usage of digital slide scanners and validation of digital pathology for routine diagnosis^13^, vast amount of raw pixel data are generated which could be interrogated to yield information beyond a simple diagnosis. Digital pathology image data is ripe ground for “data hungry” deep learning methods, which have made significant advances in the areas of image analysis, computer vision, speech recognition and speech translation, to name only a few.

In histopathology, deep learning based algorithms have been shown to detect metastatic tumor in sentinel lymph nodes of breast cancer^14^, predict mutation status in lung cancer^15^ and predict microsatellite instability (MSI)^4^, consensus molecular subtypes^18^ and outcome^17^ in CRC. More recently, Kather *et al*.^18^ extended their proposed computational pipeline^4^ to detect clinically actionable genetic alterations from H&E images. Echle *et al*.^12^ recently evaluated four models and showed that three of those models achieved a strong performance in intra-cohort validation on three big international training sets (n=1770, n=2013, and n=2197), in addition to a multi-centric dataset of The Cancer Genome Atlas (TCGA, n=426). However, both intra- and inter-cohort validations on the multi-centric TCGA dataset could only match the previous state-of-the-art performance^4^ (AUROC=0.77).

Weakly supervised learning of slide-level labels for whole-slide images (WSIs) of H&E stained tissue slides and detection of clinically actionable genetic alterations from the analysis of H&E WSIs have been shown to be possible using deep learning. However, two fundamental questions remain to be answered: 1) can we employ deep learning to predict the status of molecular pathways and relevant genetic mutations from images of H&E stained sections with a high degree of accuracy, and 2) can these predictions be mapped onto known or novel histological features?

The labeling of a digitized WSI as positive or negative without resorting to detailed annotations at cellular and regional levels would be termed as ‘weakly supervised’ learning in the language of machine learning. Recently, several weakly supervised learning methods for computational pathology have been proposed to maximize the use of available high-level slide labels.^19^ For instance, Campanella *et al*.^20^ proposed a multiple instance learning (MIL) based deep learning system to discriminate between tumor and non-tumor WSIs using high-level image labels only. However, it was shown that the MIL method Campanella *et al*.^20^ required tens of thousands of WSIs for an effective training of the deep learning model. Although the authors chose only a few representative top-ranked tiles from each slide, their method compares all tiles in the slide in each training epoch, which may be computationally expensive and requires a large amount of data for training. Another weakly supervised WSI classification method is proposed by Wang *et al*.^21^, which requires pixel-level coarse annotations that are labor-intensive or often not available as in our case, in addition to the high-level image labels. Both these methods used an additional machine learning model for aggregating tile scores into a slide score.

In this study, we propose a novel weakly-supervised deep learning pipeline which can effectively predict the status of slide labels in general and key molecular pathways and specific mutations in particular from H&E images with only a slide-level label required for training the algorithm. The proposed pipeline predicts molecular status with no cell-level or regional-level annotations of the CRC WSIs. It employs a novel strategy for sampling, which we term as the iterative draw and rank sampling (IDaRS), to train a deep learning model and efficiently identify representative image tiles (or patches) from a given WSI for a specific prediction task.

The research also relates to systematically analyzing the most discriminative visual fields in a data-driven manner to establish histologically meaningful features of each slide group in the histology landscape. In the existing literature, the histological correlates of mostly MSI are only subjectively performed, the details of which can be found in.^7,10,22–27^ IDaRS can identify the most representative tiles in each WSI for the analysis of cellular composition and TILs in each molecular pathway. To the best of our knowledge, this is the first systematic approach for analyzing the histological features associated with CRC molecular pathways and key mutations in a data-driven manner.

## Materials and methods

We propose a novel deep learning framework involving three separate models, as shown by the illustration in Fig 1. IDaRS (CNN-2, Fig 1b) is at the core of the proposed framework, which predicts the slide label (e.g., status of HM, MSI, CIN, CIMP, BRAF, and TP53) for each WSI. Since tumor tissue blocks taken during tumor sampling are large and almost always contain non-tumor tissue, it is necessary to identify the tumor areas. A fine-tuned ResNet18^28^ is used as the tumor detection model (CNN-1, Fig 1a) to extract tumor tiles from tissue contents in a given slide. These tiles serve as input to IDaRS, which learns the discriminative features of slide groups, molecular pathways and mutations for calculating a digital score of the corresponding molecular status of the WSI and its most representative visual fields. The final network (CNN-3, HoVer-Net^29^ in our case, Fig 1d) uses top-ranked tiles as predicted by IDaRS to segment and classify different types of cell nuclei in each tile for the analysis of cellular composition. Finally, the predicted digital scores of each pathway are investigated for their association with the status of CRC molecular pathways and key mutations.

**Fig 1.**
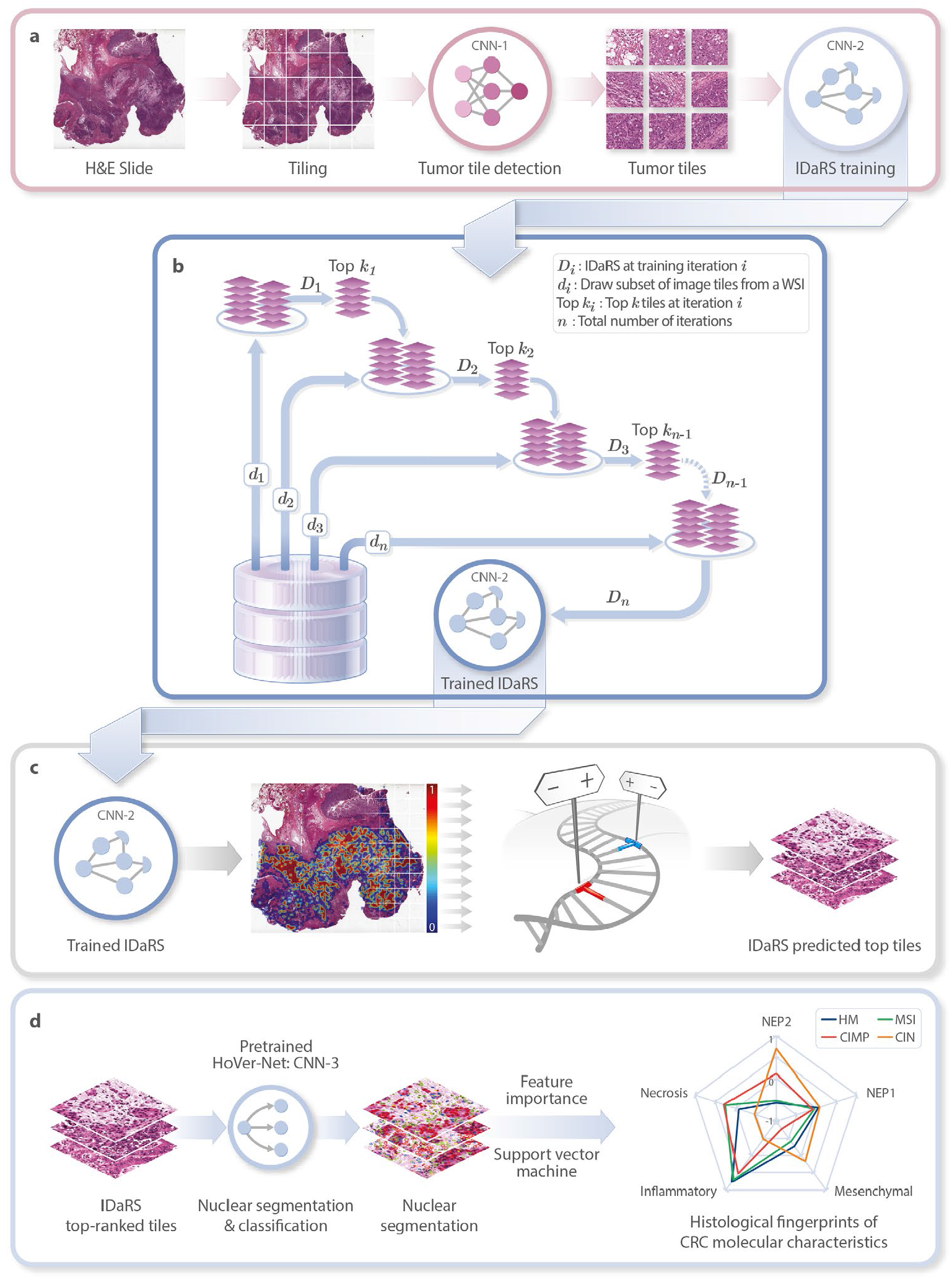
IDaRS pipeline of prediction and histopathological feature discovery of CRC pathways. **a**. Tissue segmentation and tile extraction are performed to obtain informative tiles. First deep neural network (CNN-1) is ResNet18^28^ trained on a multi-cohort dataset of CRC tissue tiles to separate tumor from non-tumor tiles and the IDaRS network model (CNN-2), a significant adaptation of ResNet34^28^, is only trained on tumor tiles for genomic instability and alteration prediction using IDaRS. **b**. The concept diagram of IDaRS illustrates a training strategy for fast labeling of WSI. The deep learning model is being trained iteratively (*D*_*i*_, *i* = 1, 2, …, *n*) for classification with a random draw (*d*_*i*_) of same number of tiles from each WSI and the *k* top tiles of the same slide drawn in the previous iteration. **c**. Trained IDaRS gives a prediction score to every tile in the WSI which are used to obtain a slide score and top-ranked tiles from each slide. **d**. Inference of HoVer-Net (CNN-3) is used to segment and classify different types of nuclei in top-ranked representative tiles for cellular composition analysis of CRC pathways. Histological patterns of CRC molecular characteristics are shown as a spider plot based on the feature importance of differential cellular composition modeled using support vector machine.

Both training and inference are used for the first two networks whereas for the third, only inference is made using a pre-trained model^i^. In the following sections, we describe the datasets used in this study and provide a detailed description of data preparation, IDaRS and histological feature discovery steps of the proposed approach.

### Data collection and preparation

The first cohort used in this study is the multi-centric cohort consisting of 502 diagnostic slides (DX) of 499 patients taken from The Cancer Genome Atlas (TCGA) colon and rectal cancer (TCGA-CRC-DX) cohort which has also been used in recent studies.^4,12,18^ This cohort has slides contributed from thirty-six different centers, mimicking a challenging real-world setting.

The IDaRS deep neural network model is trained for binary classification. For instance, in the case of HM tumor prediction, the two classes are high mutation density (HMD) and low mutation density (LMD). For MSI pathway prediction, the two classes are MSI-High (MSI) and Microsatellite stable (MSS) tumors; MSI-Low are combined with MSS in line with general literature as well as another recent study.^4^ For CIN pathway, the two classes are CIN and GS, and for CIMP pathway, the two classes are CIMP-High (CIMP-H from hereon) and CIMP-Low (CIMP-L from hereon), whereas for BRAF and TP53, the two classes are mutant (MUT) and Wild Type (WT). During the IDaRS training, MSI, CIN, CIMP-H and MUT labels are used as positive class labels and MSS, GS, CIMP-L, and WT as negative class labels.

For MSI prediction, a total of 428 WSIs (*n*=423, where *n* denotes the number of unique patients) were available with MSI labels in the TCGA-CRC-DX cohort also used in recent studies.^4,12^. The full cohort of 502 TCGA-CRC-DX WSIs is also used for BRAF and TP53 prediction whereas a subset (for which the pathway labels are available) is used for the prediction of HM, CIN and CIMP pathways. Table 1 lists the number of patients and WSIs in TCGA-CRC-DX, after excluding slides with missing metadata and those with less than ten tumor tiles, and the number of patients in different stage groupings for each pathway.

**Table 1.** Number of cases (*n*) and slides (*n*_*slides*_) in TCGA-CRC-DX and associated statistics for different CRC stages.

| | $n$ | $n_{slides}$ | Stage I | Stage II | Stage III | Stage IV |
| --- | --- | --- | --- | --- | --- | --- |
| HM/CIN | 425 | 430 | 72 | 149 | 135 | 72 |
| MSI | 423 | 428 | 72 | 149 | 135 | 72 |
| CIMP | 235 | 239 | 48 | 92 | 72 | 27 |
| BRAF & TP53 | 497 | 502 | 72 | 151 | 135 | 144 |

The ground truth labels of TCGA-CRC-DX for HMD/LMD, MSI/MSS, CIN/GS, and CIMP-H/L were obtained from Liu et al.^1^ HMD tumors are defined as those with mutation density > 10 per megabase. MSI pathways were defined as those arising from defective DNA mismatch repair. CIN exhibited marked aneuploidy defined by a clonal deletion score (> 0.0249) and GS lacked such aneuploidy. The high and low frequencies of DNA hypermethylation were used to stratify CIMP-H and CIMP-L respectively, where more than 30% of the CIMP-H tumors lacked *MLH1* silencing and MSI. The Venn diagrams shown in Fig 2 illustrate the overlap of MSI cases with other pathways (Fig 2a) and HM with MSI pathway (Fig 2b) in terms of shared number of cases.

**Fig 2.**
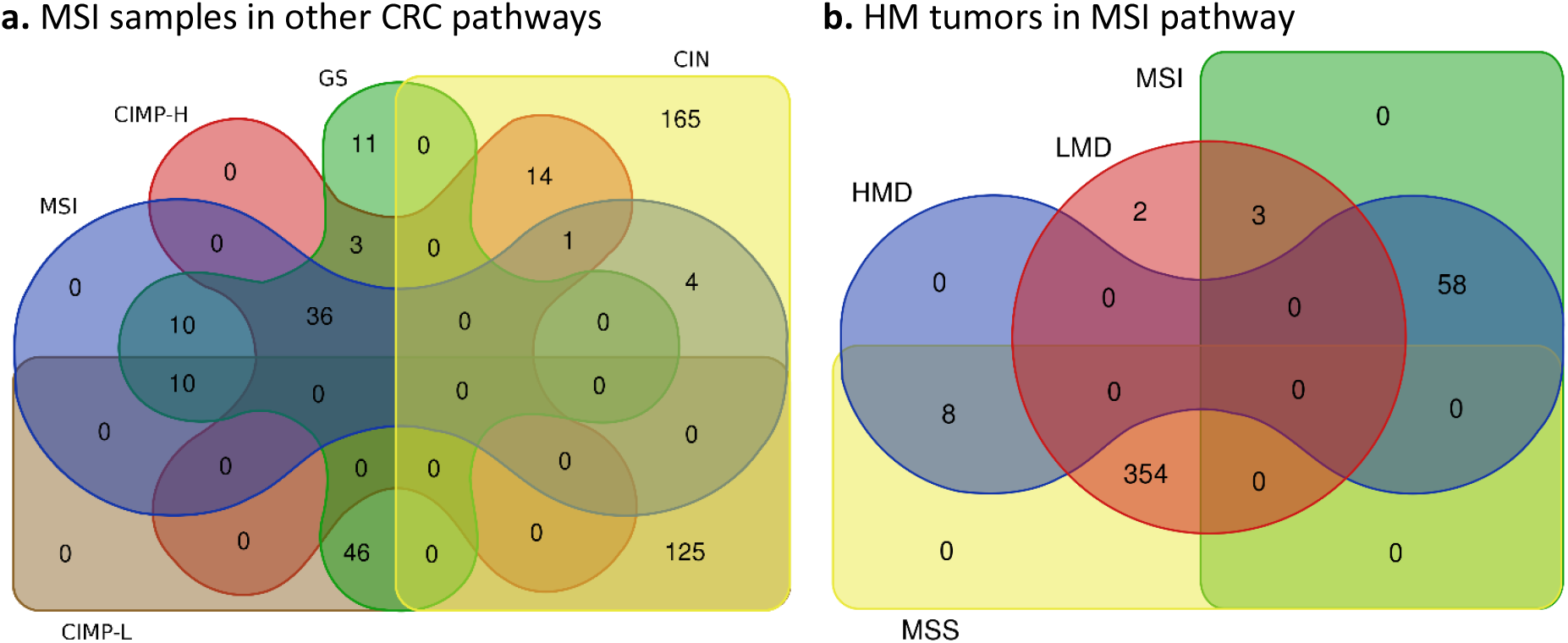
Number of samples shared among different molecular types of CRC. **a**. MSI samples in CIMP-H, GS, CIN, and CIMP-L subgroups. **b**. HM tumors in MSI and MSS subgroups.

All slides are preprocessed for tissue segmentation using Otsu thresholding.^30^ Then tiles are extracted from the segmented tissue region only. Following the protocol outlined in^31^ and used in^4,18^, tissue region in each WSI is divided into tiles of size 512×512, i.e. square tiles of 256 µm edge length at 20× magnification. Slides with less than ten tiles (three slides in total) were excluded in our experiments. For the subsequent downstream analysis through deep learning models, these tile images were resized to 1.14µm/pixel into square tiles of size 224×224. Before, training the IDaRS model, the color distribution of all tiles is normalized using structure preserving^32^ stain normalization.

### Tumor segmentation

To perform the subsequent analysis in tumor regions of the slides, a deep neural network is trained to extract tumor tiles. These tiles are taken from seven randomly picked TCGA slides and two publicly available datasets.^4,33^ Seventy percent of the data is used for training, fifteen percent for validation to fine tune for the best model and the remaining fifteen percent is held out to test the network performance with an unseen set of tiles. The model achieved an accuracy of 99% for unseen tiles. This model is then used to extract tumor tiles (a total of nearly 450K tiles) from the entire TCGA-CRC-DX cohort.

### Draw and rank sampling

A major challenge in computational pathology lies in discovering and interpreting unknown patterns from the large amount of pixel data in WSIs, especially when only a slide-level label is provided. A typical WSI may contain 150,000×100,000 pixels or more, which is too large to feed into most deep neural networks directly. Therefore, a WSI is usually divided into square tiles (or image patches) before applying deep learning, resulting in a set of tiles with limited visual context. However, the available ground truth is often a high-level label of the WSI instead of well-informed annotations at the level of tissue or cellular regions. A WSI usually consists of tens of thousands of tiles, many with no relationship with the slide-level label, where a significant and meaningful pattern may comprise a small visual field, a tile, or a few of these. It is non-trivial to decide which tiles/patches in each WSI are to be used for training. In addition, having no specific regions annotated at the cellular level is another challenge for effective training of a model.

Conventionally, a deep neural network is trained on all tiles of the training set^4^, assuming the ground truth is strongly labeled at the tile level. This is not expedient with only a slide-level label available, as there may be redundant and irrelevant tiles in the WSI resulting in a less than optimal training of the model. Besides, each WSI contributes different number of tiles, between ten to five thousand in our case, potentially introducing huge training bias to particular samples. A random selection of tiles from each WSI may solve the training bias and inefficiency but may not improve the prediction accuracy. In fact, selecting tiles that are most predictive of a given WSI label in a brute force manner is a computationally intractable problem.

In order to reduce the undesirable impact of the aforementioned issues on training, the learning problem of training a deep neural network model *f*(.; *θ*) with trainable weights *θ* can be modeled as a weakly-supervised machine learning problem with the following empirical error formulation:

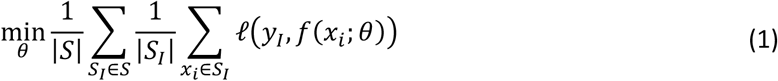

Here, *S* denotes the WSI training set with *S*_*I*_ ∈ *S* representing the set of tiles *x*_*i*_ ∈ *S*_*I*_ in the tumor region of a given WSI *I* with WSI-level label *y*_*I*_ ∈ {−1, +1}. This formulation allows choosing the most representative sub-images from each slide by minimizing the loss function *ℓ*(*y*_*I*_, *f*(*x*_*i*_; *θ*)) between the slide level label *y*_*I*_ and the tile-level output *f*(*x*_*i*_; *θ*). However, as mentioned above, calculating the loss over all tiles in a given WSI is computationally intractable. Consequently, we propose a draw-and-rank strategy that samples a set of random tiles in a given WSI using the decision function *f*(*x*_*i*_; *θ*) while maintaining the most representative tiles in an iterative manner across the training epochs. Fig 1b shows the concept diagram of the proposed IDaRS method. A more detailed description of IDaRS training is given in Algorithm 1.

The proposed algorithm uses a quasi Monte-Carlo sampling method for selection of tumor tiles from each slide in each training iteration. Specifically, for a given WSI, a representative subset of tumor tiles is obtained based on a minimum loss criterion. This also helps cope with the information density and training bias because of the varied number of image tiles from each WSI. Since random tiles are chosen in a training set, which may or may not contain discriminative tiles in each iteration, each training iteration concludes by carrying the top *k* tiles from the current iteration as a part of the set of most predictive tiles through to the next iteration. We choose top *k* tiles from each slide ranked by the maximum probability of a tile for every class label. The top *k* predicted tiles of each slide are iteratively learned and assumed to be the most representative tiles from the slide.

#### Algorithm 1 IDaRS training algorithm.

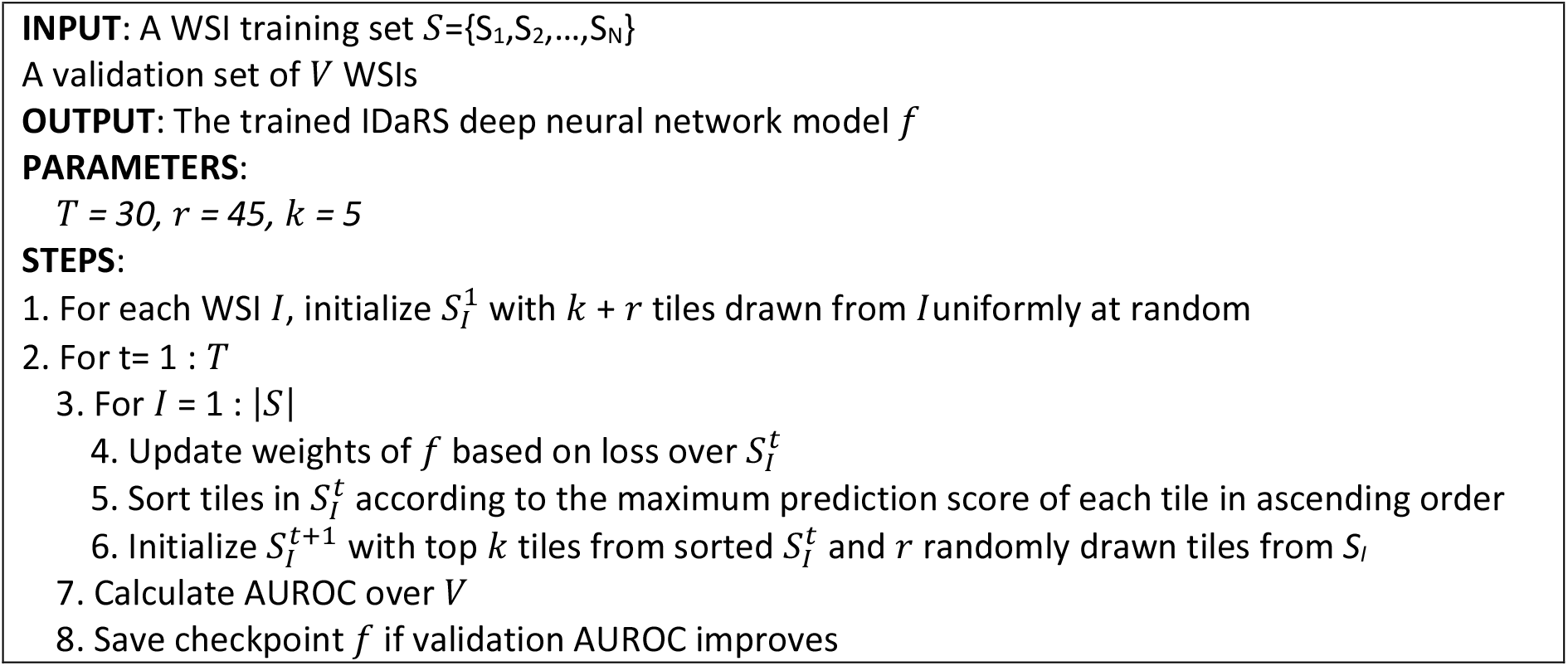

### Neural architecture and training

Our neural network model is a fine-tuned version of ResNet34^28^, pre-trained on ImageNet, for the prediction of slide labels, i.e. the status of molecular pathway and genetic mutations. Training is performed for 30 iterations with a batch size of 256. In each iteration, the top 5 tiles are carried to the next training iteration along with 45 randomly selected tiles from each slide. Thus, a training subset of only 50 image tiles from each slide was selected for each iteration.

We use the symmetric cross entropy (SCE) loss^34^ during the training process to overcome the risk of training error associated with weak labels, mathematically shown as *ℓ*_*SCE*_ = *αℓ_CE_* + *βℓ_RCE_*. The values of *α* and *β* are found empirically (*α* = 2 and *β* = 3) by trying different combinations between 0 and 5, as reported in original research.^34^ The SCE loss *ℓ*_*SCE*_ is a weighted sum of standard CE *ℓ*_*CE*_ and reversed cross entropy *ℓ*_*RCE*_ losses. The SCE loss enforces a balance between learning and robustness to weakly labeled tile samples.

The deep learning model trained using IDaRS gives a probability score to each tile. This score is meant to be a likelihood of a tile belonging to the positive class in the binary classification setting. Scores of all tiles in a WSI are considered for aggregating into a WSI score and the results are reported for average probability based aggregation after comparing different aggregation schemes – which included majority vote, average top ten probabilities in a WSI, maximum of probabilities, average of probabilities, geometric mean of probabilities, median of probabilities, and average of the probabilities which are greater than the median probability in a WSI – to obtain the best aggregation scheme for each prediction problem.

The training procedure was guided by an Adam optimizer^35^ with learning rate and weight decay of 0.4×10^−4^. The PyTorch deep learning library^36^ was used for the implementation. A set of data augmentation including random rotation with maximum angle of 15 degrees, random horizontal and vertical flip transforms available in PyTorch are applied on-the-fly. Additional data augmentations of center-crop, random resized crop, and random crop of image size 224×224 are used on all input tiles (of size 256×256) to get multiple of it centered at different spatial locations in the tile. A validation set is used for saving checkpoints as an early stopping criterion during the model training. All experiments are performed on Nvidia DGX-2 Deep Learning System with 16× 32GB Tesla V100 Volta GPUs in a shared environment. The deep learning model is built on a single GPU equipped with dedicated RAM of 32 GB and 6 worker threads.

### Histological feature discovery of CRC pathways

IDaRS offers opportunities of objective quantitative analyses of differential histological features of CRC pathways at cellular levels in the H&E histology landscape in a systematic manner. We have analyzed nuclear composition of IDaRS-predicted top tiles for each pathway and HM tumors. This is determined independently by the nucleus segmentation and classification network HoVer-Net^29^, also termed as *CNN-3* in Fig 1c. This step mimics the visual analysis of an expert pathologist who analyzes nuclear composition in the tissue microenvironment subjectively by looking on the slide under the microscope at high power field. We use a deep neural network trained on a publicly available dataset for the analysis of tumor microenvironment and nuclei types to enable an objective and systematic analysis of the cellular composition.

For this purpose, we first performed nucleus segmentation of the top predicted tiles using HoVer-Net^29^, a recent state-of-the-art nuclear segmentation and classification model. We employed the HoVer-Net model pre-trained on the PanNuke dataset^37^ for nucleus segmentation and classification. The model processed each of the top predictive IDaRS tiles to analyze their nuclear composition by segmenting and classifying tile nuclei into one of the following five categories: neoplastic and non-neoplastic epithelial, inflammatory, mesenchymal, and dead cells. The cell segmentation and classification results of HoVer-Net were visually examined and agreed by expert pathologists for all cell types. Non-neoplastic epithelial cells identified by the network were found to be different from those identified as neoplastic cells, but all of those were categorized as tumor cells by the experts.

This was as expected, since only the tumor tiles were used for the training and inference of IDaRS. Therefore, neoplastic cells identified by HoVer-Net are termed as neoplastic epithelial type 1 (NEP1) and non-neoplastic epithelial identified by the network are termed as neoplastic epithelial type 2 (NEP2) cells.

Keeping in view the so-called “black box” challenge of deep learning, the cellular composition analysis also enables us to interpret the IDaRS predictions and helps us to better understand the different slide groups and molecular pathways objectively.

### Performance evaluation and comparison

We use the area under the convex hull of the receiver operating characteristic (AUROC) as performance measures of the model predictions. The comparison is made with the state-of-the-art automated MSI prediction results as a benchmark problem on the TCGA-CRC-DX cohort with exactly the same train/test splits^4^, multi-fold cross validation for MSI^12^, and other pathways and mutations (BRAF and TP53).^18^

### Existing state-of-the-art results

To the best of our knowledge, Kather *et al*.^4^ were the first to propose a deep learning based automated prediction of MSI status from H&E images. They use the same deep learning pipeline with more data and multiple cohorts in^12^ to predict MSI status from H&E images. The authors used standard fully supervised image classification by transfer learning the ResNet18 model^28^ pre-trained on ImageNet. Standard CE loss was used to train the model with Adam optimizer with learning rate and weight decay of 0.4×10^−4^. The training dataset of color normalized tiles was balanced by under sampling of the majority class (MSS). Tile scores obtained from the deep learning models are aggregated into slide score using average or majority voting. Again to the best of our knowledge, the study by Kather et al.^18^ was the first that reports automatic CIN and HM tumors as well as BRAF and TP53 prediction results, which considered mutation prediction as a fully supervised deep learning problem. To overcome the bias resulting from some WSIs having a large number of tiles, each WSI was allowed to contribute a maximum of 1,000 tiles. No color normalization method was used. The *Shufflenet*^38^ model pre-trained on ImageNet was used for transfer learning, with the CE loss and an Adam optimizer with learning rate and weight decay of 0.4×10^−4^.

## Results

The deep learning pipeline for clinical pathways and mutation prediction is developed and validated using the TCGA-CRC-DX cohort with similar experimental setups to those reported in published studies.^4,12,18^ The results from applying our IDaRS based digital scores for the prediction of HM, MSI, CIN, CIMP, BRAF and TP53 status from H&E slides are shown in Table 2 and Table 3. For HM prediction, only 67 HMD and 363 LMD labeled patients are considered for binary classification, and IDaRS yields an average cross-validation AUROC of 0.81±0.04.

**Table 2.** AUROC based comparative analysis of HM, MSI, CIN, CIMP, BRAF and TP53 on TCGA-CRC-DX using 4-fold cross validation. In the second column, labels mentioned first (HMD, MSI, CIN, CIMP-H, MUT) are considered as positive (Pos) and those mentioned next (LMD, MSS, GS, CIMP-L, WT) are considered as negative (Neg). The next column (Published AUROC) lists the current state-of-the-art results^12,18^ of 3-fold cross validation average AUROC reported for the same cohort. The last column (IDaRS AUROC) shows the result of 4-fold cross validated average AUROC produced using the proposed IDaRS algorithm.

| Predicting | No. of Samples<br>(Pos, Neg) | Published AUROC | IDaRS<br>AUROC |
| --- | --- | --- | --- |
| HM | 430 (HMD: 67, LMD: 363) | 0.71<sup>18</sup> | 0.81 |
| MSI | 428 (MSI: 62, MSS: 366) | 0.74<sup>12</sup> | 0.86 |
| CIN | 430 (CIN: 313, GS: 117) | 0.73<sup>18</sup> | 0.83 |
| CIMP | 239 (CIMP-H:55, CIMP-L: 184) | - | 0.79 |
| BRAF | 502 (MUT: 59, WT: 443) | 0.66<sup>18</sup> | 0.79 |
| TP53 | 502 (MUT: 294, WT: 208) | 0.64<sup>18</sup> | 0.73 |

**Table 3.** IDaRS internal and external validation of MSI prediction on the TCGA and PAIP datasets.

| Problem | No. of Samples (Pos, Neg) | Published AUROC | AUROC | AUPRC |
| --- | --- | --- | --- | --- |
| MSI – TCGA (internal) | MSI: 62, MSS: 298 | 0.77<sup>4</sup> | 0.90 | 0.72 |
| MSI - External | MSI: 12, MSS: 35 | - | 0.98 | 0.95 |

TCGA-CRC-DX used in^12^ contains 62 cases of MSI and 366 cases of MSI-Low or MSS. The IDaRS method with an average cross-validated AUROC of 0.86±0.04 significantly outperforms the published AUROC of 0.74.^12^ We used 4-fold cross validation for all experiments reported in Table 2 to measure the average IDaRS performance on the entire cohort in an internal cross-validation manner. We used two folds for training, one fold as a validation set for keeping the best performing model and another one held-out for measuring the performance of the model on an unseen test set. In case there were multiple slides of the same patient, we ensure that they were grouped in the same fold.

The proposed algorithm uses a quasi Monte-Carlo sampling method for selection of tumor tiles from each slide in each training iteration. Specifically, for a given WSI, a representative subset of tumor tiles is obtained based on the minimum loss criterion. This also helps cope with the information density and training bias because of the varied number of image tiles from each WSI.

Since tiles are randomly chosen in a training set, which may or may not contain discriminative tiles in each iteration, each training iteration concludes by carrying the top *k* tiles from the current iteration forward into the set of most predictive tiles for the next iteration. The top *k* predicted tiles of each slide are assumed to be the most discriminative tiles from the slide for prediction of the slide label.

Considering the stochastic element of IDaRS, each fold is executed three times to get average AUROC and standard deviation per fold, which is further averaged over each fold to get average AUROC and standard deviation of multi-fold cross validation. The corresponding AUROC curves are graphically shown in Fig 3. For CIN, 313 patients labeled as CIN and 117 as GS are used for binary classification. The IDaRS method with an average cross-validation AUROC of 0.83±0.02 has significantly outperformed the published AUROC of 0.73.^18^ For CIMP, only 55 CIMP-H and 184 CIMP-L labeled patients are considered for binary classification. We also predict HMD/LMD and CIMP-H/CIMP-L status using IDaRS with an average AUROC of 0.81±0.04 and 0.79±0.05, respectively.

**Fig 3.**
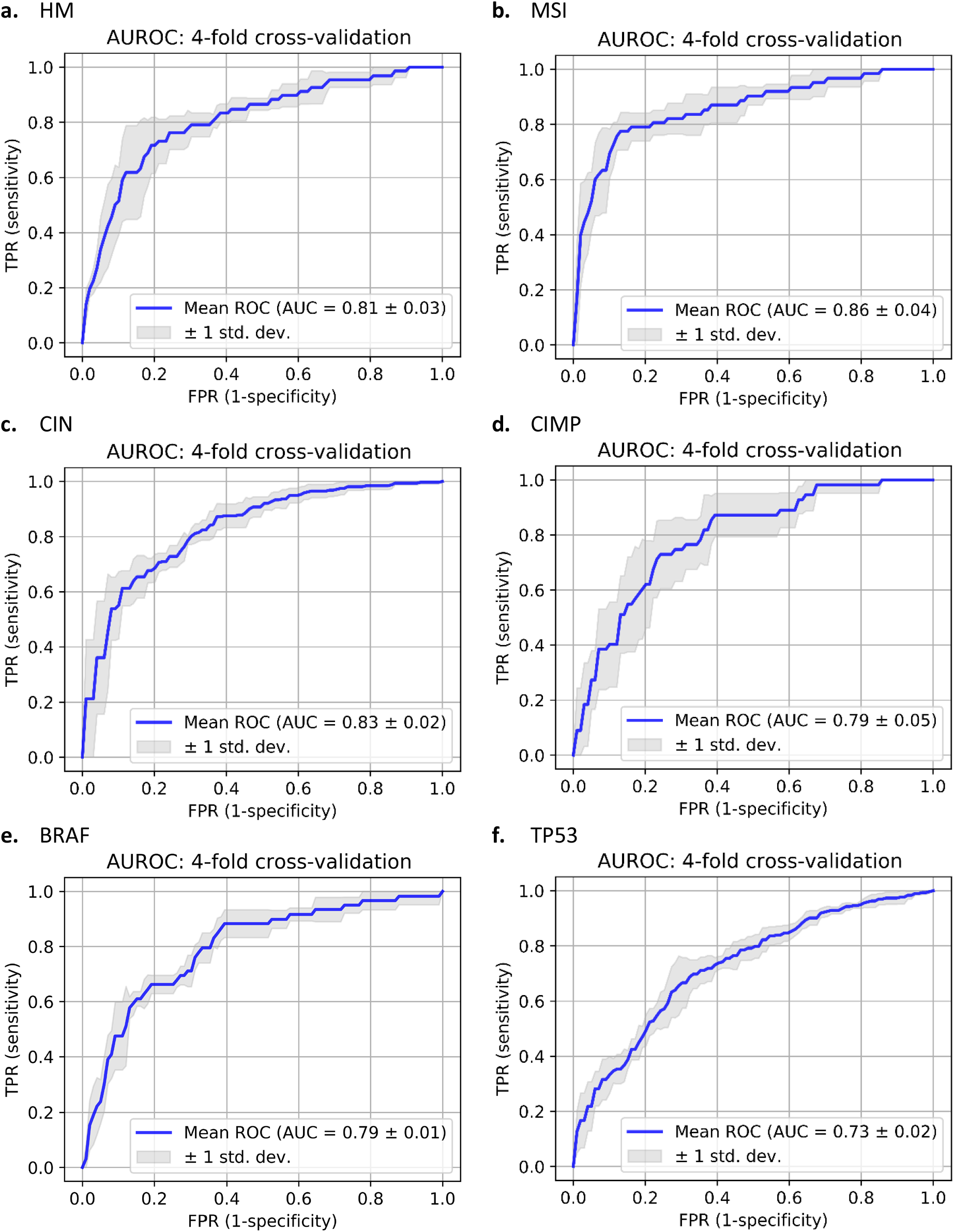
IDaRS based prediction of CRC pathways on TCGA-CRC-DX. AUROC plots of cross validation average AUROC **a-f**. For each curve, *y*-axis is true positive rate (TPR) (sensitivity) and *x*-axis false positive rate (FPR) (1-specificity). **a**. HM, **b**. MSI, **c**. CIN, **d**. CIMP, **e**. BRAF and **f**. TP53.

The IDaRS method with an average AUROC of 0.79 for BRAF and 0.73 for TP53 has significantly outperformed the published AUROC values of 0.66 for BRAF and 0.64 for TP53. All resulting ROC curves for four-fold cross validation, to ascertain the generalizability of the proposed method, are shown in Fig 3, with shaded areas covering the variation in the ROC curves for the different folds. Considering varying ratio of class imbalance in reported experiments, we also computed the average precision of precision-recall curve (AUPRC). AUPRC is also averaged for multiple runs and multiple folds of the experiments. The highest AUPRC is obtained for the prediction of CIN, with an acceptable range for MSI, TP53 and CIMP (see Supplementary Materials).

### CRC pathway prediction in tumors grouped into different stages

Clinical staging of CRC, as in many other cancers, is used to establish the extent of disease spread to help in determining suitable treatment strategies. Knowing the status of MSI is important for the choice of adjuvant chemotherapy and immunotherapy for CRC.^40^ Immunotherapy is usually recommended as the first-line treatment for early stage MSI CRCs ^39^ and second-line treatment for stage IV^41^ MSI CRCs due to evidence for its significantly better disease-free survival and overall survival. Patients with stage III MSI-L/MSS CRC have better overall survival with adjuvant chemotherapy.^41^ CIN and BRAF mutation have been associated with poor overall survival while the jury is still out for CIMP.^2,3,6,7,9,10,26^ We consider the accurate prediction of MSI status for CRC tumors to be a strength of IDaRS based digital MSI scores.

Considering this importance of clinical stage in routine histopathology and CRC pathways and mutations, we divided the entire TCGA-CRC-DX cohort into four groups of patients, one group per stage, and computed AUROC for each stage group separately. We used the IDaRS scores obtained using the models trained in cross-validation, for which the results are reported in Table 2. In Fig 4, we plot the AUROC of corresponding molecular type, pathway, and mutation prediction for four different stage groups. A stagewise breakdown of the cases is provided in Table 1. The prediction performance of same type, pathway, and mutation varies for different stages. Moreover, the number of cases and class ratio also varies for each pathway and mutation in each stage. Among the individual stage groups, HM, CIN, and BRAF are predicted with the highest AUROC in stage IV, whereas digital IDaRS based MSI score gives the highest accuracy for stage I and II. Prediction accuracy of the MSI status for Stage I and II groups is higher than that for Stage III and IV groups. For Stage IV cases, our method produces the highest HM, CIN, and BRAF prediction accuracy of 0.98, 0.94, and 0.90, respectively. CIMP-H and CIMP-L are much better differentiable by our method for stage I to stage III cases than for stage IV cases.

**Fig 4.**
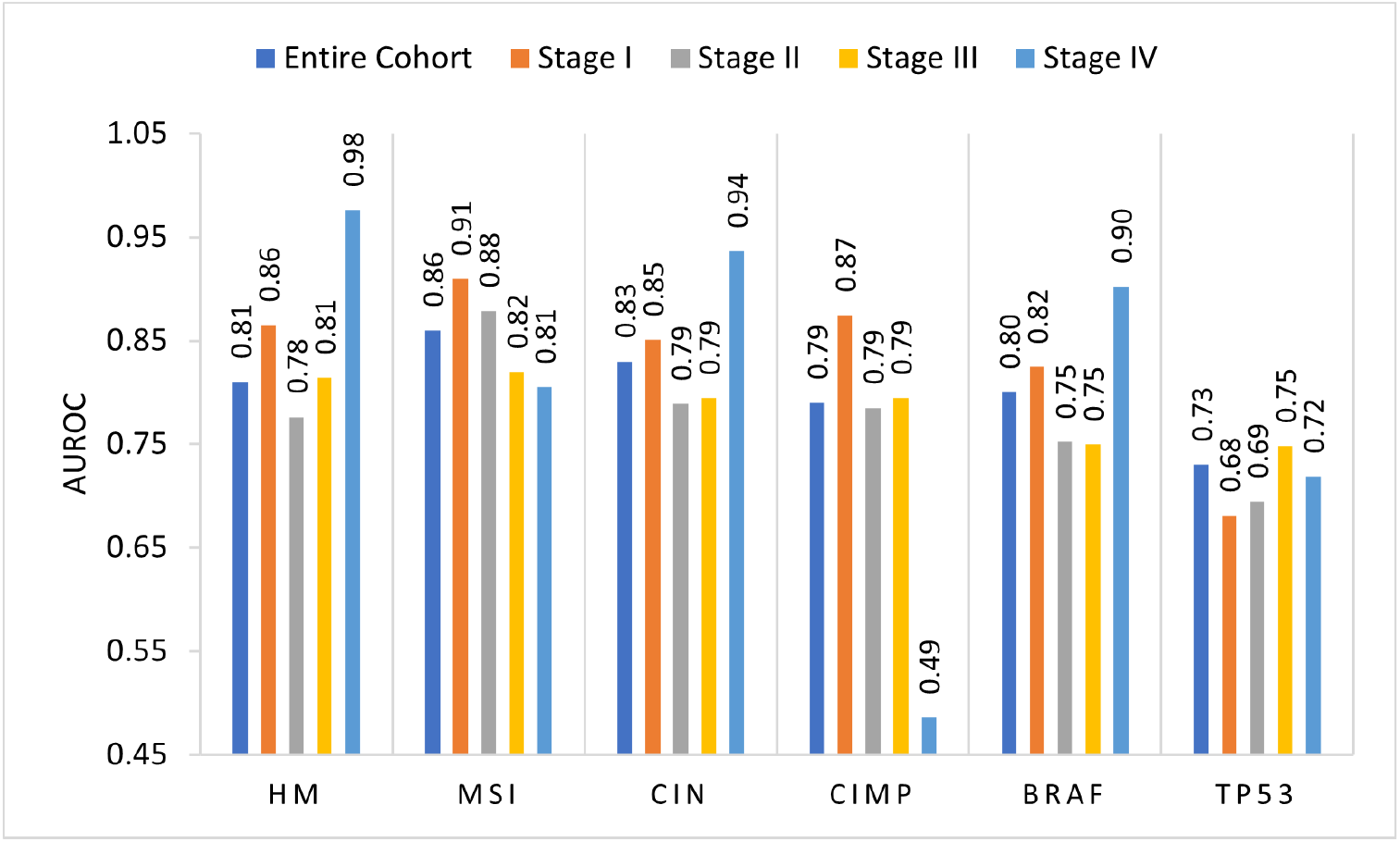
IDaRS predictions for pathways and mutation different stage groups. AUROC on *y*-axis and pathways and mutations on the *x*-axis. Entire cohort refers to the whole TCGA-CRC-DX cohort considering patients of all stages as one group. For each stage, AUROC is computed for a subgroup of patients belonging to the same stage while leaving out the patients belonging to other stages.

### Internal and external validation for MSI prediction

The train-test split experiment for MSI prediction reported in Kather et al.^4^ is an older version of TCGA-CRC-DX, which contained 62 cases of MSI and 298 cases of MSS. For a fair comparison, we have used the same patient cohort and train/test splits for MSI prediction. The IDaRS method with an average AUROC of 0.90 significantly outperforms the previous state-of-the-art AUROC of 0.77 by Kather et al. For MSI-TCGA (internal) results in Table 3, both the training and testing splits are from TCGA-CRC-DX cohort. For MSI-External test set, the IDaRS model was trained on TCGA-CRC-DX cohort and tested on the PAIP challenge cohort, which contains 47 slides in total (12 MSI and 35 MSS) from three different centers. The tumor tiles (58,097 in total) are obtained from PAIP cohort using the expert pathologist annotations of the tumor region provided together with the PAIP challenge dataset.

### Differential cellular composition of CRC pathways

We used a feature analysis approach to understand the association between cellular composition of top-ranked tiles generated by IDaRS for a given WSI and different CRC pathways and HM tumors.

Table 4 shows the internal train-test split experiments on the prediction of HM, MSI, CIN and CIMP pathways. Further analysis of the cellular composition is performed on the top ten most predictive tiles in each WSI of the unseen test set (99 WSIs) as predicted by the IDaRS algorithm.

**Table 4.** IDaRS evaluation on internal test set used for cellular composition analysis.

| Predicting | No. of Samples (Positive, Negative) | IDaRS AUROC | IDaRS AUPRC |
| --- | --- | --- | --- |
| HM | HMD: 66, LMD: 293 | 0.88 | 0.66 |
| MSI | MSI: 62, MSS: 297 | 0.90 | 0.72 |
| CIN | CIN: 257, GS: 102 | 0.85 | 0.92 |
| CIMP | CIMP-H: 51, CIMP-L: 152 | 0.84 | 0.61 |

For cellular composition analysis, we first employed in-house nucleus segmentation and classification method HoVer-Net^29^ to localize, segment and classify different types of cells in a tile into NEP1, NEP2, inflammatory, mesenchymal, and necrosis as shown for some example images in Fig 5. We then used the counts of individual cell types in the tile, henceforth termed as its *cellular composition profile*, as features to differentiate between most predictive tiles from corresponding different CRC pathways (MSI *vs* MSS, CIMP-High *vs* CIMP-Low and CIN *vs* GS) and HM tumors (HMD *vs* LMD) through four separate linear Support Vector Machine (SVM) predictors. For illustration, few examples of IDaRS predicted most representative tiles for each molecular label are shown in Fig 6.

**Fig 5.**
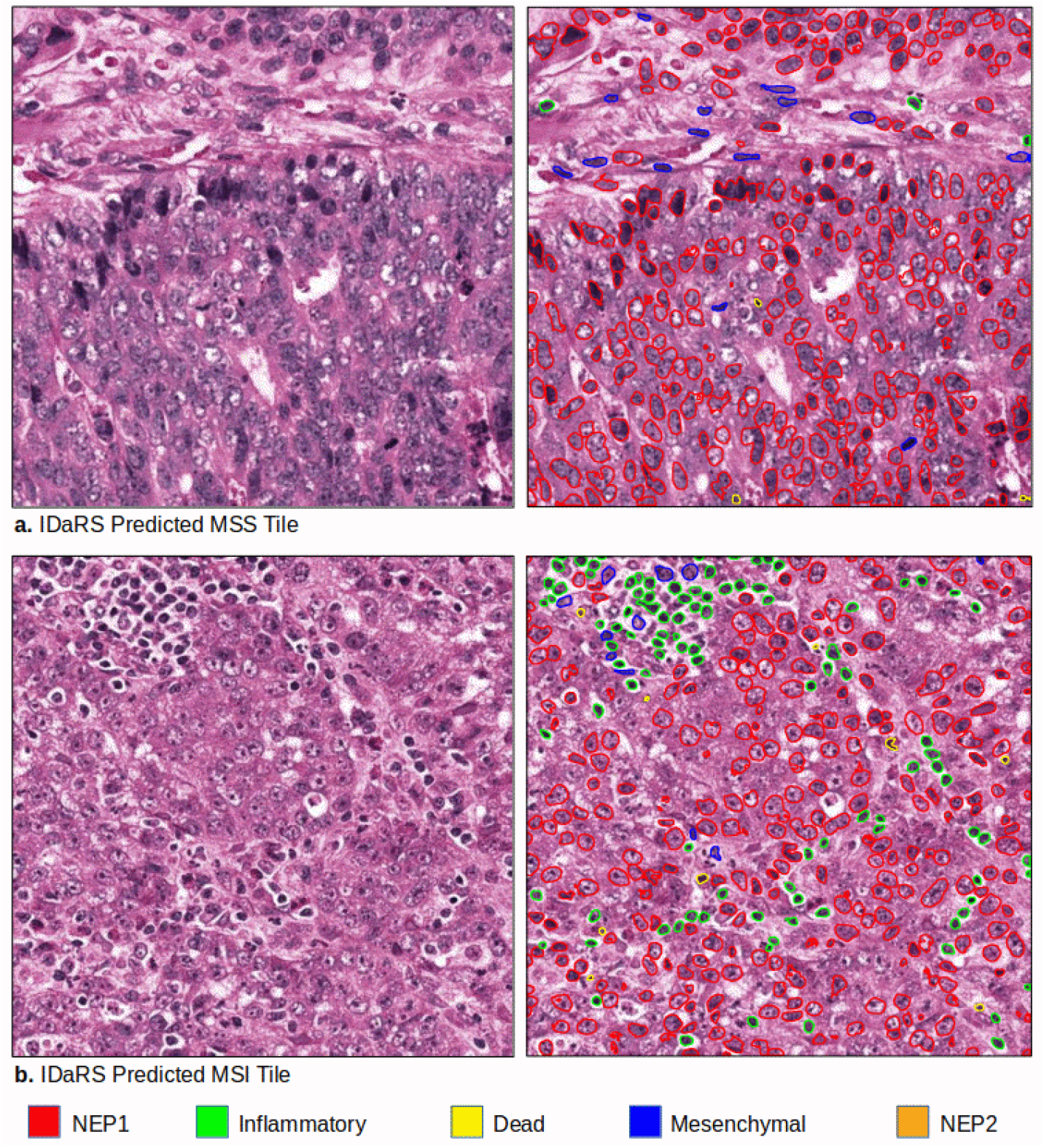
Nuclear segmentation and classification by HoVer-Net. Examples of **a**. top predicted MSS tile (left) with its HoVer-Net output (right) and **b**. top predicted MSI tile (left) with its HoVer-Net output (left) for tumor microenvironment analysis. Detected nuclei of different types are circled with different colors: red: NEP1, green: inflammatory, blue: mesenchymal, yellow: necrotic, and orange: NEP2.

**Fig 6.**
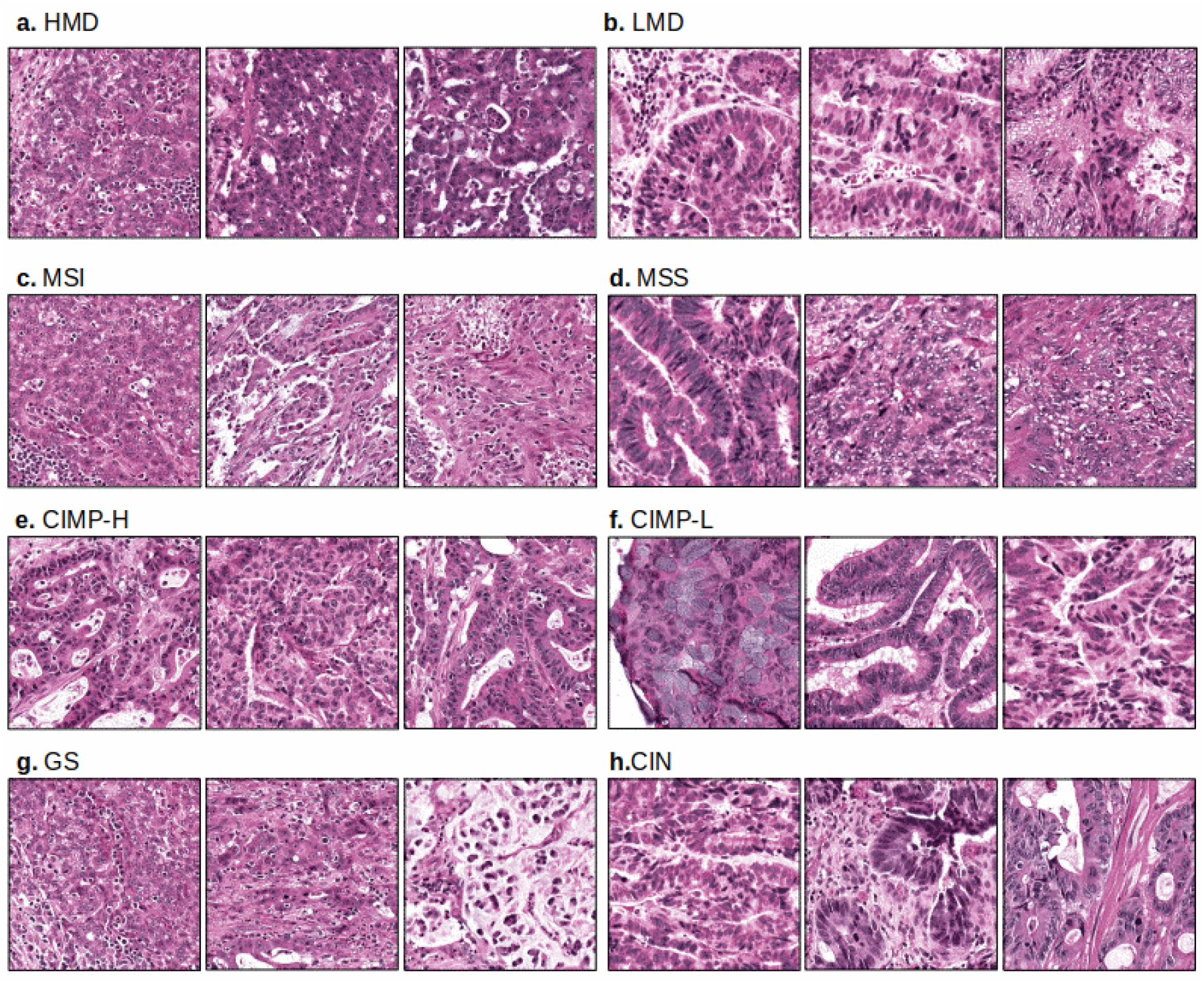
IDaRS discovered most representative visual fields. Examples of IDaRS predicted visual fields from different slides associated to **a**. HMD, **b**. LMD, **c**. MSI, **d**. MSS, **e**. CIMP-H, **f**. CIMP-L, **g**. GS, and **h**. CIN classes.

The weight vector of a linear SVM allows an estimation of the relative importance of different cell types in predictions of the CRC pathways.^42–44^

Specifically, we analyzed the bootstrap averages of SVM weights across 100 runs over top tiles to get estimates of their relative contribution to the prediction of CRC pathways. The magnitude of different components of the resulting weight vectors as an indication of the feature importance, as shown in Fig 7. for the four molecular classification problems, can be interpreted as the relative degree to which the over- or under- representation of different types of cells (NEP 1, NEP 2, inflammatory, mesenchymal, and dead) in the top ten tiles is predictive of the corresponding class label (HMD vs. LMD, MSI vs. MSS, CIMP-High vs. Low and CIN vs. GS).

**Fig 7.**
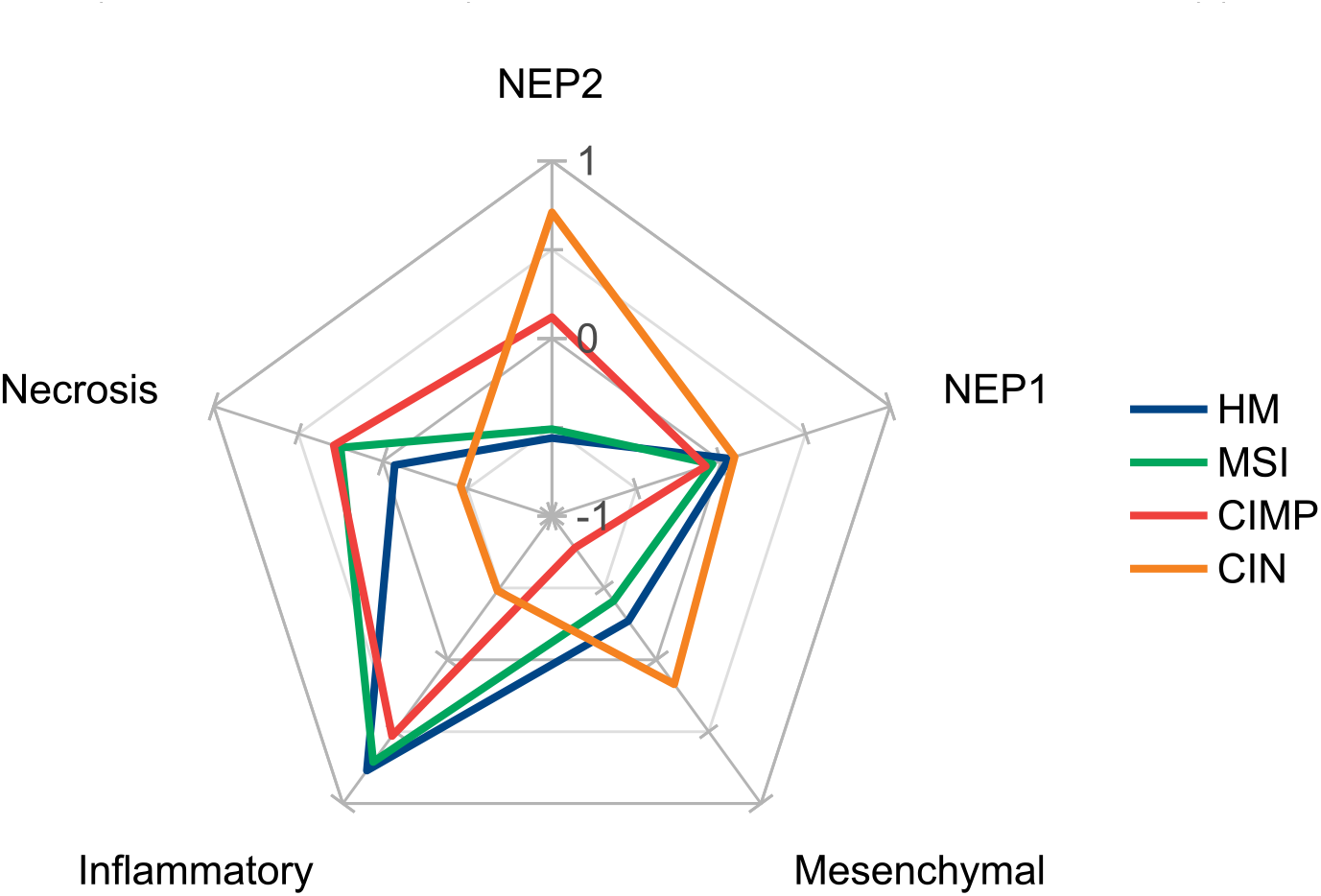
Spider chart represents differential cellular composition as histological fingerprint of CRC molecular pathways and HM tumors. Normalized weight between −1 and 1 show the magnitude of significance of the corresponding histological feature for different molecular characteristics (HM, MSI, CIMP and CIN). Radii represent cell types (necrosis, NEP1, mesenchymal, inflammatory and NEP2) as a differential histological feature. Points of a given molecular label on each radii when connected forms a fingerprint for corresponding molecular type. Relatively high proportion of inflammatory and necrotic cells is differential feature of HM, MSI, and CIMP-H cases. Relatively high magnitude of NEP2 and necrotic cells for CIMP-H and relatively low magnitude of necrosis and high magnitude of mesenchymal for HMD, separate CIMP-H and HMD from MSI, respectively. Relatively high proportion of NEP1 and NEP2, and mesenchymal cells are differential features of CIN.

The understanding of CRC stratification into multiple pathways, their morphological correlates and clinical significance is still evolving and most findings so far are inconclusive. For instance, Gao *et al*.^23^ have associated MSI with low inflammatory infiltration but not with the absence of tumor necrosis whereas other studies have found tumor infiltrating lymphocytes^22,10^ as a strong predictor of MSI with lack of tumor necrosis. In another recent study, Saller *et al*.^45^ confirmed three cases showing similar immunogenic response, but inconsistent interpretation of their MSI status on the review with different MSI assays. This contradiction may be attributed to the lack of an objective and systematic data-driven approach for analyzing the entire tissue slide for multi-centric cohorts. We believe IDaRS based analysis presented here is a first step towards an objective, quantitative and reproducible approach for analyzing CRC pathways from routine histology slides and study their corresponding histological correlates in a systematic and data-driven manner.

Fig 7 shows that relatively high proportion of inflammatory and tumor necrosis (dead cells) and a relatively low proportion of neoplastic epithelial (both NEP1 and NEP2) and mesenchymal cells are associated with MSI. Relatively high proportion of inflammatory and NEP1 cells and relatively low proportion of NEP2, mesenchymal and tumor necrotic cells are associated with HM. Similarly, relatively high proportion of inflammatory, tumor necrosis, and NEP1, and a relatively low proportion of mesenchymal cells and NEP2 are associated with CIMP-H. In contrast, high proportion of NEP2, mesenchymal, and NEP1 cells and relatively low proportion of tumor necrosis and inflammatory cells are associated with CIN. A strong association of CIN subgroup with the presence of NEP2, mesenchymal and NEP1 cells and the absence of necrosis and inflammatory cells may explain its association with a less favorable outcome in patients, as has been suggested in the literature^2,10^. A strong association of CIMP-H with the presence of inflammatory and the absence of mesenchymal cells may explain its relationship with molecular characteristics of MSI as an active immune response as reported previously.^26^ These findings also suggest that MSI is strongly associated with the presence of inflammatory cells and with the absence of NEP1 cells and moderately associated with tumor necrosis and absence of mesenchymal and NEP2 cells. This may explain the relatively more favorable response of MSI tumor to the immunotherapy.^40,41^

The prognostic significance of TILs in general and its association with MSI has already been established in existing literature.^24,25^ Fig 7 also suggests infiltration by inflammatory cells as a key discriminating histological feature in MSI and HM tumors. Therefore, we quantified TILs of the two subclasses in each pathway and HM tumors graphically shown in Fig 8 computed from the top ten IDaRS tiles per WSI using a TIL abundance score.^46^ A paired t-test found TIL scores to be statistically significant for the MSI pathway (p<0.0024) and HM tumors (p<0.024) but not for the CIMP (p<0.09) and the CIN (p<0.157) pathways.

**Fig 8.**
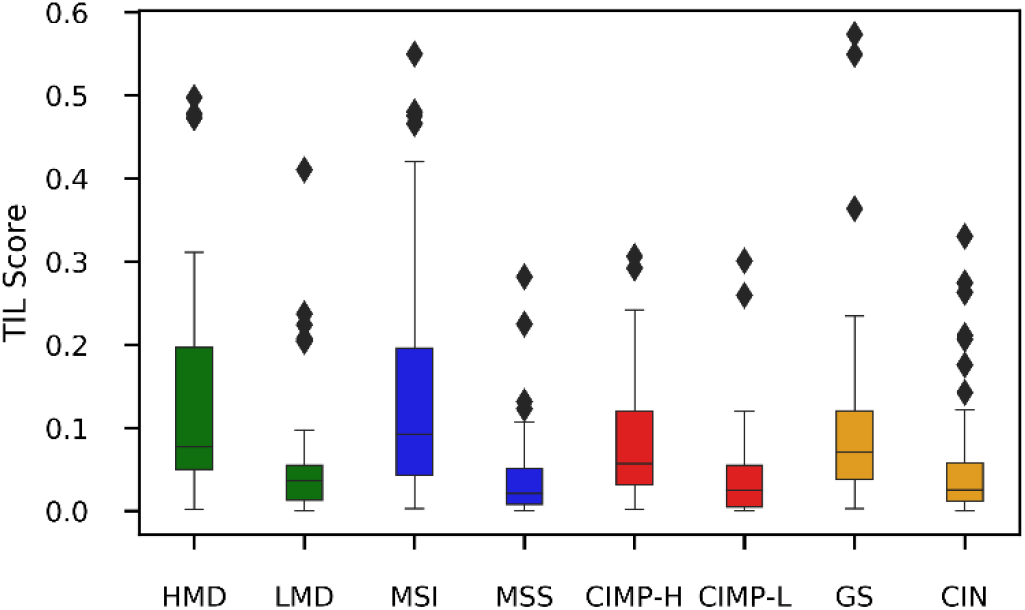
Quantifying TILs in HMD, MSI, CIMP-H, GS, MSS, CIMP-L and CIN. TIL abundance score (*y*-axis) and CRC pathways (*x*-axis). In IDaRS predicted top ten tiles, HMD and MSI samples got the highest TIL score. CIMP-H and GS sample also got higher TIL scores than CIMP-L and CIN.

### Correlation analysis of the digital IDaRS based scores

Here, we analyze the relationship of HM tumors and three CRC pathways in terms of which tumor microenvironment is activated for predicting different molecular label. We trained three deep learning models separately using the same patient-wise splits of training, validation and testing datasets. Each patient has multiple slide-level ground truths of being HMD/LMD, MSI/MSS, CIN/GS, and CIMP-H/CIMP-L available. A heatmap of one test set sample is shown in Fig 9 for the visual illustration of different types of the microenvironments active in different molecular labels. The IDaRS predicted top ten tiles for each label (HMD, MSI, GS, and CIMP-H) are different showing different kind of microenvironment is activated in the same image in response to different WSI label.

**Fig 9.**
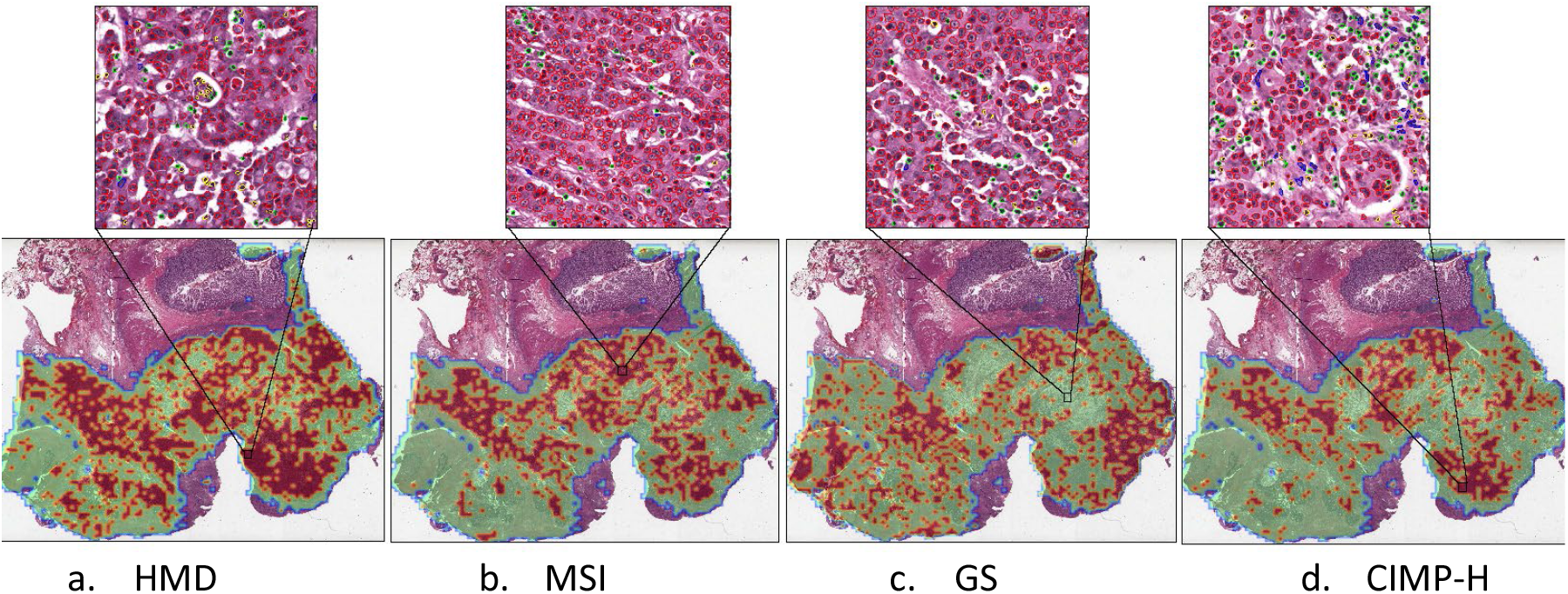
IDaRS tile prediction heatmaps-1. A test set slide labeled as HMD, MSI, GS and CIMP-H, WT for BRAF, TP53 and KRAS mutations. IDaRS assigned tile probabilities of being (HMD, MSI, CIN, CIMP-H) are used to generate a heatmap overlay. In the heatmap, the red color corresponds to the positive class and green to the negative class; **a**. Overlay heatmap of HMD prediction, HMD: Red. **b**. Overlay heatmap of MSI prediction, MSI: Red. **c**. Overlay heatmap of CIN prediction, GS: Green. **d**. Overlay heatmap of CIMP-H prediction, CIMP-H: Red. An IDaRS predicted top tile is also shown for each label.

A heatmap of another test set sample is shown in Fig 10 for visual illustration. It is observed that all the ground truth labels LMD, MSS, CIN, and CIMP-L mostly activate in the same region of the slide but not at tile level, all top ten tiles associated to each label are different. For each pathway, both positive and negative tiles are active in the slide, however, tiles associated to the ground truth label are fewer in number than other labels. This indicates, more sophisticated methods (e.g. another machine learning method) of aggregation than simple average and majority voting might be favorable to correctly classify this case. Moreover, this could be linked to the difficulty of MSI scoring especially when the immunogenic response of an MSS identified cases is similar to the MSI histomorphology.^45^

**Fig 10.**
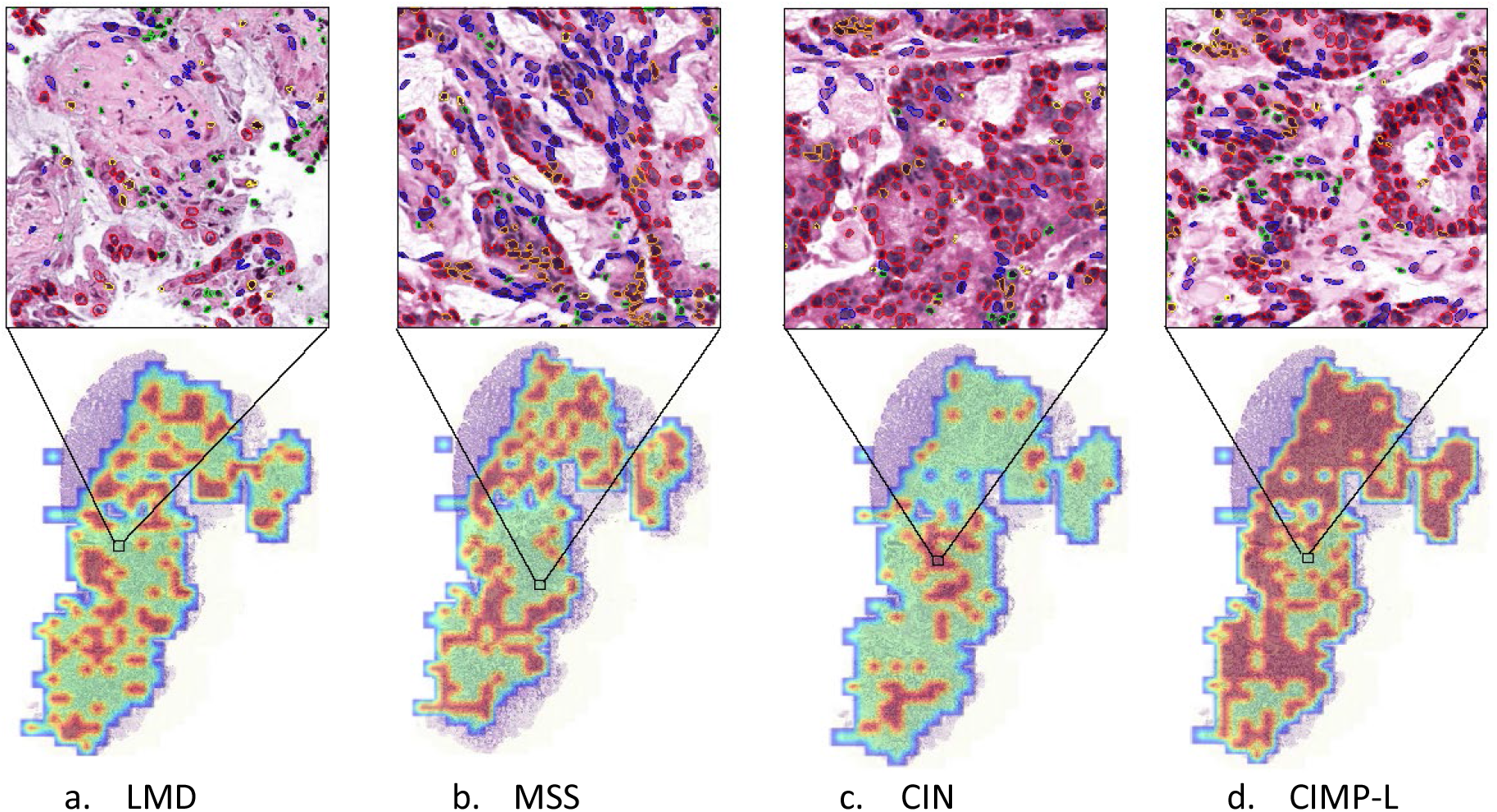
IDaRS tile prediction heatmaps-2. A test set slide with known KRAS mutation and wild type BRAF showing IDaRS derived overlay heat map labeled as LMD, MSS, CIN and CIMP-L. In the heatmap, the red color corresponds to the positive class and green to the negative class; **a**. Overlay heatmap of LMD prediction, LMD: Green. **b**. Overlay heatmap of MSS prediction, MSS: Green. **c**. overlay heatmap of CIN prediction, CIN: Red. **d**. overlay heatmap of CIMP-L prediction, CIMP-L: Green. An IDaRS predicted top tile is also shown for each label.

We conducted an investigation into correlating the IDaRS based digital scores of HM, MSI, CIN, CIMP, BRAF and TP53 for each slide of the TCGA-CRC-DX test set used in^4^ obtained by applying separately trained models on the same training set. Fig 11a and Fig 11b show scatterplots of our IDaRS based digital scores for HM and MSI, and HM and CIMP pathways, respectively. These plots demonstrate strong positive correlation (using the Pearson correlation coefficient *r*) between the digital scores of HM and MSI (*r*=0.81, *p*<10^−23^) and also between the digital scores of HM and CIMP (*r*=0.65, *p*<10^−12^). Fig 11c shows the scatterplot of our IDaRS based digital scores for MSI and CIMP, again demonstrating strong positive correlation between the two digital scores (*r*=0.68, *p*<10^−14^) in line with existing literature^7,10,26^ and showing that most of the MSI samples are also predicted as CIMP-H and MSS as CIMP-L. Among the single labels, MSS shows highest positive correlation of 0.96 (*p*<10^−40^) with CIMP-L and MSI shows positive correlation of 0.49 with CIMP-H (*p*<0.07). Our digital scores of MSI and CIN also show strong negative correlation (*r*=−0.75, *p*<10^−18^) though it did not necessarily endorse mutual exclusivity for all MSI with CIN cases. Most MSS cases and few MSI cases also got high CIN score, which is in line with previous findings about MSI and CIN (and as can also be observed in the Venn diagrams of Fig 2).^2,5^

**Fig 11.**
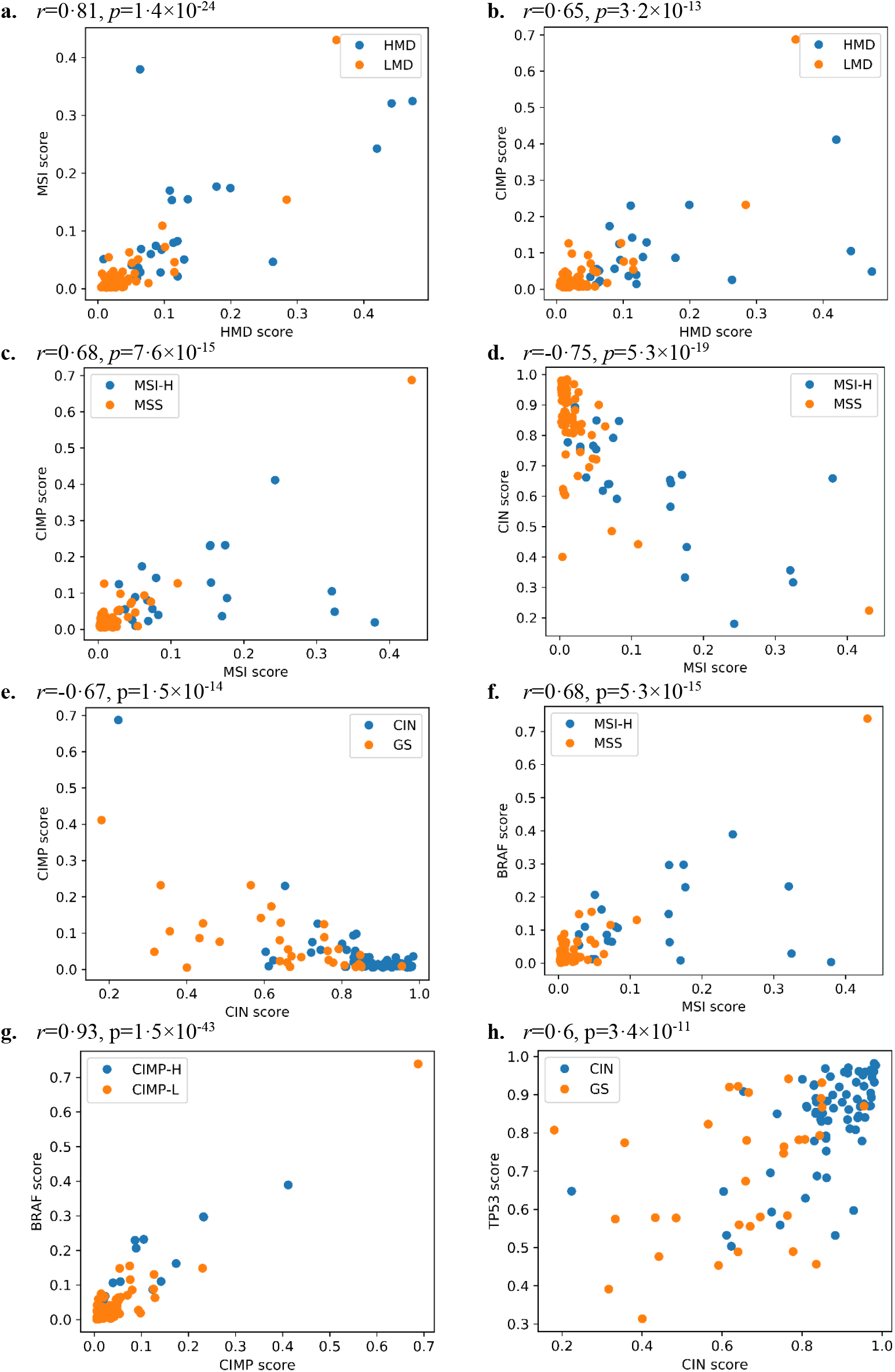
Correlations in pathways and mutations. Digital IDaRS scores for HM, MSI, CIN, CIMP, BRAF and TP53 of each slide of the test set obtained by applying separately trained models on the training set of the same slides: **a**. HMD and MSI, positively correlated. **b**. HMD and CIMP, positively correlated. **a**.**b**. Points are colored using HM ground truth labels. **c**. MSI and CIMP, positively correlated. IDaRS predicted MSI score on x-axis and CIMP score on y-axis. **d**. MSI and CIN, negatively correlated. MSI score on x-axis and CIN score on y-axis. **c**.**d**. Points are colored using MSI ground truth labels. **e**. CIMP and CIN, negatively correlated. MSI score on x-axis and CIN score on y-axis. Points are colored using CIN ground truth labels. **f**. MSI and BRAF, positively correlated. MSI score on x-axis and BRAF score on y-axis. Points are colored using MSI or MSS ground truth label. **g**. CIMP and BRAF, positively correlated. CIMP score on x-axis and BRAF score on y-axis. Points are colored using CIMP ground truth labels. **h**. CIN and TP53, positively correlated. CIN score on x-axis and TP53 score on y-axis. Points are colored using CIN ground truth labels.

The IDaRS based digital scores of CIN and CIMP also show negative correlation (*r*=−0.67, *p*<10^−13^), with high CIN scoring cases being scored low in CIMP and with overlapping GS cases. MSI have highest negative correlation of −0.68 (*p*<0.0005) with GS whereas CIMP-L has negative correlation of −0.76 with CIN (*p*<10^−12^). IDaRS predictions also suggest MSI and CIMP are positively correlated to BRAF mutation with correlation coefficients of 0.68 (*p*<10^−14^) and 0.93 (*p*<10^−42^) whereas CIN has highest positive correlation of 0.60 (*p*<10^−10^) to TP53 mutation.

## Discussion

In this study, we have shown that by employing a novel training strategy in a standard deep learning model can help achieve a significant improvement in the prediction of CRC HM tumors, molecular pathways and genetic mutations. We optimized our tumor segmentation algorithm, which ensures that the normal colon mucosa is not included in the analysis.

Our extensive experimentation leads us to believe that key CRC molecular pathways and genetic mutations are predictable from digitized images of H&E stained whole slides with higher accuracy than the state-of-the-art methods. HM, MSI, CIN, CIMP, BRAF, and TP53 are strongly predictable from CRC tumors in H&E images than whole-genome duplication, KRAS, PIK3CA, SMAD4, SMAD2, and APC – our method achieved a performance of AUROC < 0.7 for these mutations, which we do not consider to be significant.

Our analysis of the stage-wise prediction accuracy (AUROC) indicates that prediction of HM tumors, molecular pathways (MSI, CIN and CIMP) and key mutations (BRAF and TP53) may vary between stage groupings. For example, MSI prediction dropped slightly from Stage I (AUROC=0.91) to Stage IV (AUROC=0.81). This indicates that tumor stage may affect deep learning based discrimination of predictive features. Since we interrogated sections of the primary tumors only, it may be that the deep features of MSI cancers at an early stage are subtly different from those of advanced MSI cancers. This may be due to increased heterogeneity of the advanced MSI tumors. Similarly, the prediction accuracies of HM tumors, CIN pathway and BRAF mutation are the highest in Stage IV (AUROC=0.98, 0.94 and 0.90, respectively). We argue that it may be worth exploring the predictive value of deep features in early stage cancers before they become advanced stage. If so, these features may allow more aggressive treatment of some early stage CRC cases that have a risk of disease progression.

One key challenge for MSI prediction is the tradeoff between sensitivity and specificity of the clinical gold-standard testing in addition to inconsistencies among different MSI testing assays. Further improvements in AUROC of MSI and comparison with other molecular and genetic labels are possible if the specificity and sensitivity of the corresponding testing used for acquiring the slide level ground truth labels are known beforehand.

We explored the cellular composition analysis as a way of mining significant differential features for each of the CRC pathways. We found tumor-lymphocyte and tumor-necrosis infiltrates are associated with HM, MSI, CIMP-H, and GS groups. Some of these findings are in line with the existing literature and produce new knowledge about each pathway concerning different cell types, which requires large-scale validation and further exploration with subsequent studies. Doing this analysis in a systematic data-driven manner in this study is a significant and innovative aspect of this study in addition to the robustness of its findings of differential cellular composition. The differential cellular composition analysis suggests that infiltration scores of lymphocytes and necrosis within the tumor can serve as a digital biomarker for further stratification and analysis of each pathway and its clinical impact.

## Conclusions

We proposed an iterative draw-and-rank sampling (IDaRS) strategy for fast prediction of the slide label in general, and the status of hypermutated tumors, molecular pathways and genetic mutations in particular, in digitized whole-slide images of routine H&E stained tissue slides. We showed that the proposed method produces the state-of-the-art performance in terms of AUROC for automatically predicting the status of molecular subtypes (HM, MSI, CIN, CIMP) and key mutations (BRAF and TP53) in WSIs of CRC samples from the TCGA and another independent cohort. The histological patterns and features mined from the strongly predicted tiles were analyzed and found to be consistent with existing findings about the HM tumors and CRC pathways.

We found that relatively high proportion of inflammatory and necrosis with relatively low proportion of neoplastic and mesenchymal cells appeared in CIMP-High and MSI cases, which may explain why these cases are more likely respond positively to immunotherapy. Increased neoplastic, necrotic, and mesenchymal cells with decreased inflammatory cells may be attributed to less favorable patient outcome, as evidenced in the CIN phenotype, which has been previously linked with low rates of overall and progression-free survival.

Automatic identification of genetic pathways (especially MSI) from ubiquitously available H&E images carries significant potential to provide time and cost-effective decision support to improve patient management and care by efficiently identifying patients who could benefit from immunotherapy and targeted therapy. It further presents an opportunity to objectively verify existing knowledge and produce new knowledge about visual pathology and patient outcomes associated with different pathways of CRC. Further large-scale validation of these findings is required for their implementation in diagnostic practice and patient management.

## Data Availability

All images and the associated pathways/mutations status for the TCGA cohort (COAD and READ) used in this study are publicly available at https://portal.gdc.cancer.gov/ and cbioportal. A link to the TCGA manifest file that can be used to download all images for the TCGA cohort can be found in the Supplementary Materials document. The ground truth labels of TCGA-CRC-DX for HMD/LMD, MSI/MSS, CIN/GS, and CIMP-H/L were obtained from Liu et al. A link to the spreadsheet containing the corresponding clinical and molecular data including cancer stages, subtypes and the status of mutations and pathways can also be found in the Supplementary Materials. De-identified pathology images and annotations from Pathology AI Platform (PAIP) used with institutional permissions in this study can be obtained via appropriate data access requests through the URL: http://www.wisepaip.org/paip.

http://www.wisepaip.org/paip

## Acknowledgments

The research reported in this publication was supported by the UK Medical Research Council (award MR/P015476/1).

The results shown here are in whole or in part based upon data generated by the TCGA Research Network [https://www.cancer.gov/tcga].

De-identified pathology images and annotations in the PAIP cohort (used as external validation cohort in this study) were prepared and provided by the Seoul National University Hospital by a grant of the Korea Health Technology R&D Project through the Korea Health Industry Development Institute (KHIDI), funded by the Ministry of Health & Welfare, Republic of Korea (grant number: HI18C0316).

## Contributors

MB and NR conceived the study. FM and SR contributed to study design. MB, FM, SR, and NR contributed to the methodology. NR was responsible for the overall project supervision. NR and MB coordinated the study. MB provided informatics support and conducted the experiments. MB, FM, and NR contributed to the literature search. MB contributed to data collection. AA contributed to data curation. MB, SG, FM, SR and NR contributed to data analysis. MB, SR, FM and NR contributed to the figure design. IC, MI, DS, AA, and NR contributed to expert review. All authors contributed to data interpretation. MB prepared the initial draft. All authors contributed to the writing, review, or revision of the manuscript.

## Limitations

The proposed IDaRS algorithm uses Monte Carlo sampling to iteratively select training data from each WSI, leading to a slightly different trained model for each run. Therefore, we have run each experiment three times with the same data splits and averaged measures are reported in the results. The data imbalance is another common issue with the medical datasets, which can affect the classification performance. Keeping this in view, we have reported the AUPRC results as well the AUROC results in the Supplementary Materials. External validation is done on 47 WSIs of the PAIP dataset obtained from three different centers, which is a relatively small dataset. A pretrained HoVer-Net model is used for nuclei segmentation and classification followed by a review by expert pathologists (AA and DS). Some example images and their corresponding results for nuclear segmentation and classification are provided in the Supplementary Materials document. Although the TCGA cohort is a multi-centric dataset and PAIP is an external cohort, further large-scale validation of these findings is required before their implementation in diagnostic practice and patient management.

## Data Sharing

All images and the associated pathways/mutations status for the TCGA cohort (COAD and READ) used in this study are publicly available at https://portal.gdc.cancer.gov/ and cbioportal. A link to the TCGA manifest file that can be used to download all images for the TCGA cohort can be found in the Supplementary Materials document. The ground truth labels of TCGA-CRC-DX for HMD/LMD, MSI/MSS, CIN/GS, and CIMP-H/L were obtained from Liu et al.^1^ A link to the spreadsheet containing the corresponding clinical and molecular data including cancer stages, subtypes and the status of mutations and pathways can also be found in the Supplementary Materials. De-identified pathology images and annotations from Pathology AI Platform (PAIP) used with institutional permissions in this study can be obtained via appropriate data access requests through the URL: http://www.wisepaip.org/paip.

## Footnotes

i https://github.com/simongraham/hovernet_inference

## References

1. Liu Y, Sethi NS, Hinoue T, Schneider BG, Cherniack AD, Sanchez-Vega F, et al. Comparative Molecular Analysis of Gastrointestinal Adenocarcinomas. Cancer Cell. 2018 Apr;33(4):721–735.e8.

2. Pino MS, Chung DC. The Chromosomal Instability Pathway in Colon Cancer. Gastroenterology. 2010;138(6):2059–72.

3. Singh MP, Rai S, Pandey A, Singh NK, Srivastava S. Molecular subtypes of colorectal cancer: An emerging therapeutic opportunity for personalized medicine. Genes Dis [Internet]. 2019; Available from: http://www.sciencedirect.com/science/article/pii/S235230421930100X

4. Kather JN, Pearson AT, Halama N, Jäger D, Krause J, Loosen SH, et al. Deep learning can predict microsatellite instability directly from histology in gastrointestinal cancer. Nat Med. 2019;25(7):1054–1056.

5. Goel A, Arnold CN, Niedzwiecki D, Chang DK, Ricciardiello L, Carethers JM, et al. Characterization of Sporadic Colon Cancer by Patterns of Genomic Instability. Cancer Res. 2003;63(7):1608–1614.

6. Al-Sohaily S, Biankin A, Leong R, Kohonen-Corish M, Warusavitarne J. Molecular pathways in colorectal cancer. J Gastroenterol Hepatol. 2012;27(9):1423–31.

7. Bae JM, Kim JH, Kang GH. Molecular Subtypes of Colorectal Cancer and Their Clinicopathologic Features, With an Emphasis on the Serrated Neoplasia Pathway. Arch Pathol Lab Med. 2016;140(5):406–12.

8. Guinney J, Dienstmann R, Wang X, de Reyniès A, Schlicker A, Soneson C, et al. The consensus molecular subtypes of colorectal cancer. Nat Med. 2015 Nov;21(11):1350–6.

9. Moreno V, Rebeca S-P. Altered pathways and colorectal cancer prognosis. BMC Med. 2015;13(76):1–3.

10. Shia J, Schultz N, Kuk D, Vakiani E, Middha S, Segal NH, et al. Morphological characterization of colorectal cancers in The Cancer Genome Atlas reveals distinct morphology–molecular associations: clinical and biological implications. Mod Pathol. 2017 Apr;30(4):599–609.

11. FDA Approves First-Line Immunotherapy for Patients with MSI-H/dMMR Metastatic Colorectal Cancer. US Food Drug Adm FDA [Internet]. 2020 Jun [cited 2020 Jan 7]; Available from: https://www.fda.gov/news-events/press-announcements/fda-approves-first-line-immunotherapy-patients-msi-hdmmr-metastatic-colorectal-cancer

12. Amelie E, Heike IG, Philip Q, et al. Clinical-grade Detection of Microsatellite Instability in Colorectal Tumors by Deep Learning. Gastroenterology [Internet]. 2020; Available from: http://www.sciencedirect.com/science/article/pii/S0016508520348186

13. Snead DR, Tsang Y-W, Meskiri A, Kimani PK, Crossman R, Rajpoot NM, et al. Validation of digital pathology imaging for primary histopathological diagnosis. Histopathology. 2016;68(7):1063–1072.

14. Bejnordi BE, Veta M, Van Diest PJ, Van Ginneken B, Karssemeijer N, Litjens G, et al. Diagnostic assessment of deep learning algorithms for detection of lymph node metastases in women with breast cancer. Jama. 2017;318(22):2199–2210.

15. Coudray N, Ocampo PS, Sakellaropoulos T, Narula N, Snuderl M, Fenyö D, et al. Classification and Mutation Prediction from Non-Small Cell Lung Cancer Histopathology Images using Deep Learning. Nat Med. 2018;24(10):1559–1567.

16. Sirinukunwattana K, Domingo E, Richman SD, Redmond KL, Blake A, Verrill C, et al. Image-based consensus molecular subtype (imCMS) classification of colorectal cancer using deep learning. Gut [Internet]. 2020; Available from: https://gut.bmj.com/content/early/2020/07/19/gutjnl-2019-319866

17. Skrede O, Raedt SD, Kleppe A, Hveem T, Danielsen H. Deep learning for prediction of colorectal cancer outcome: a discovery and validation study. The Lancet. 2020;395:350–60.

18. Kather JN, Heij LR, Grabsch HI, Loeffler C, Echle A, Muti HS, et al. Pan-cancer image-based detection of clinically actionable genetic alterations. Nat Cancer. 2020 Aug;1:789–799.

19. Rony J, Belharbi S, Dolz J, Ayed IB, McCaffrey L, Granger E. Deep weakly-supervised learning methods for classification and localization in histology images: a survey. ArXiv Prepr ArXiv190903354. 2019;

20. Campanella G, Hanna MG, Geneslaw L, Miraflor AP, Silva VWK, Busam KJ, et al. Clinical-grade computational pathology using weakly supervised deep learning on whole slide images. Nat Med. 2019;1–9.

21. Wang X, Chen H, Gan C, Lin H, Dou Q, Tsougenis E, et al. Weakly Supervised Deep Learning for Whole Slide Lung Cancer Image Analysis. IEEE Trans Cybern. 2019;1–13.

22. Greenson JK, Huang S-C, Herron C, al et. Pathologic predictors of microsatellite instability in colorectal cancer. Am J Surg Pathol. 2009;33(1):126–133.

23. Gao J-F, Arbman G, Wadhra TI, Zhang H, Sun X-F. Relationships of Tumor Inflammatory Infiltration and Necrosis With Microsatellite Instability in Colorectal Cancers. World J Gastroenterolgy. 2005;11(14):2179–2183.

24. Smyrk TC, Watson P, Kaul K, Lynch HT. Tumor-infiltrating lymphocytes are a marker for microsatellite instability in colorectal carcinoma. Cancer. 2001;91(12):2417–22.

25. Hendry S, Salgado R, Gevaert T, Russell PA, John T, Thapa B, et al. Assessing Tumor-infiltrating Lymphocytes in Solid Tumors: A Practical Review for Pathologists and Proposal for a Standardized Method from the International Immunooncology Biomarkers Working Group: Part 1: Assessing the Host Immune Response, TILs in Invasive Breast Carcinoma and Ductal Carcinoma in Situ, Metastatic Tumor Deposits and Areas for Further Research. Adv Anat Pathol. 2017;24(5):235–251.

26. Advani SM, Advani P, DeSantis SM, Brown D, VonVille HM, Lam M, et al. Clinical, Pathological, and Molecular Characteristics of CpG Island Methylator Phenotype in Colorectal Cancer: A Systematic Review and Meta-analysis. Transl Oncol. 2018;11(5):1188–201.

27. Alexander J, Watanabe T, Wu T-T, Rashid A, Li S, Hamilton S. Histopathological Identification of Colon Cancer with Microsatellite Instability. Am J Pathol. 2001;158(2):527–535.

28. He K, Zhang X, Ren S, Sun J. Deep Residual Learning for Image Recognition. In: The IEEE Conference on Computer Vision and Pattern Recognition (CVPR). 2016.

29. Graham S, Vu QD, Raza SEA, Azam A, Tsang YW, Kwak JT, et al. Hover-net: Simultaneous segmentation and classification of nuclei in multi-tissue histology images. Med Image Anal. 2019;101563.

30. Otsu N. A Threshold Selection Method from Gray-Level Histograms. N/A. 1979;9(1):62–66.

31. Muti HS, Loeffler C, Echle A, Heij LR, Buelow RD, Krause J, et al. The Aachen Protocol for Deep Learning Histopathology: A hands-on guide for data preprocessing [Internet]. 2020. Available from: https://doi.org/10.5281/zenodo.3694994

32. Vahadane A, Peng T, Sethi A, Albarqouni S, Wang L, Baust M, et al. Structure-Preserving Color Normalization and Sparse Stain Separation for Histological Images. IEEE Trans Med Imaging. 2016;35(8):1962–71.

33. Shaban M, Awan R, Fraz MM, Azam A, Tsang Y, Snead D, et al. Context-Aware Convolutional Neural Network for Grading of Colorectal Cancer Histology Images. IEEE Trans Med Imaging. 2020;1–1.

34. Wang Y, Ma X, Chen Z, Luo Y, Yi J, Bailey J. Symmetric Cross Entropy for Robust Learning with Noisy Labels. ArXiv. 2019;abs/1908.06112.

35. Kingma DP, Ba J. Adam: A Method for Stochastic Optimization. In: Bengio Y, LeCun Y, editors. 3rd International Conference on Learning Representations, ICLR 2015, San Diego, CA, USA, May 7-9, 2015, Conference Track Proceedings [Internet]. 2015. Available from: http://arxiv.org/abs/1412.6980

36. Paszke A, Gross S, Massa F, Lerer A, Bradbury J, Chanan G, et al. PyTorch: An imperative style, high-performance deep learning library. In: Advances in Neural Information Processing Systems. 2019. p. 8024–8035.

37. Gamper J, Koohbanani NA, Graham S, Jahanifar M, Benet K, Khurram SA, et al. PanNuke Dataset Extension, Insights and Baselines. ArXiv Prepr ArXiv200310778. 2020;

38. Zhang X, Zhou X, Lin M, Sun J. ShuffleNet: An Extremely Efficient Convolutional Neural Network for Mobile Devices. In: The IEEE Conference on Computer Vision and Pattern Recognition (CVPR). 2018.

39. Chalabi M, Fanchi LF, Van den Berg JG, Beets GL, Lopez-Yurda M, Aalbers AG, et al. Neoadjuvant ipilimumab plus nivolumab in early stage colon cancer. Ann Oncol. 2018 Oct;29:viii731.

40. Kang S, Na Y, Joung SY, Lee SI, Oh SC, Min BW. The significance of microsatellite instability in colorectal cancer after controlling for clinicopathological factors: Medicine (Baltimore). 2018 Mar;97(9):e0019.

41. Sun BL. Current Microsatellite Instability Testing in Management of Colorectal Cancer. Clin Colorectal Cancer. 2020 Aug;S1533002820301043.

42. Minhas F ul AA, Asif A, Arif M. CAFÉ-Map: Context Aware Feature Mapping for mining high dimensional biomedical data. Comput Biol Med. 2016;79:68–79.

43. Ben-Hur A, Ong CS, Sonnenburg S, Schölkopf B, Rätsch G. Support Vector Machines and Kernels for Computational Biology. PLOS Comput Biol. 2008;4(10):1–10.

44. Chang Y-W, Lin C-J. Feature Ranking Using Linear SVM. In: Guyon I, Aliferis C, Cooper G, Elisseeff A, Pellet J-P, Spirtes P, et al., editors. Hong Kong: PMLR; 2008. p. 53–64. (Proceedings of Machine Learning Research; vol. 3). Available from: http://proceedings.mlr.press/v3/chang08a.html

45. Saller J, Qin D, Felder S, Coppola D. Microsatellite Stable Colorectal Cancer With an Immunogenic Phenotype: Challenges in Diagnosis and Treatment. Clin Colorectal Cancer. 2020 Jun;19(2):123–31.

46. Shaban M, Khurram SA, Fraz MM, Alsubaie N, Masood I, Mushtaq S, et al. A Novel Digital Score for Abundance of Tumour Infiltrating Lymphocytes Predicts Disease Free Survival in Oral Squamous Cell Carcinoma. Sci Rep. 2019;9.

